# Emotion-Mind Dynamic (EMD): Face-to-face and online blended-learning life coaching for a social prescribing intervention to support mental health and wellbeing: Protocol for a Social Return on Investment evaluation

**DOI:** 10.1101/2022.01.24.22269523

**Authors:** M. Lynch, A Makanjuola, N. Hartfiel, A. Cuthbert, E. Winrow, R.T. Edwards

## Abstract

**Introduction:** The NHS is experiencing increased pressures due to an ageing population and rising health inequalities. GPs spend almost a fifth of their patient facing time addressing non-medical problems such as social isolation, financial struggles, and bereavement. Recent research suggests that holistic approaches linking social care to primary care may be cost-effective in supporting patients. Social Prescribing (SP) is an approach which could help individuals by sign posting and referral to non-clinical services, which can promote improvements in mental health and wellbeing, building resilience using community assets. The EmotionMind Dynamic (EMD) is a lifestyle coaching programme that supports individuals suffering from anxiety or depression referred from the health and social care sectors. EMD offers a unique, non-clinical mixed-modality approach combining coaching, mentoring, counselling skills, teaching and mindfulness. EMD is an adaptable, guided self-help tool that can be used as a preventative, supportive or reactive measure for improving mental health and wellbeing.

**Methods:** SROI methodology will be used to evaluate the EMD service. The aim of the SROI analysis is to develop a programme-level theory of change to establish how inputs (e.g. costs, staffing) are converted into outputs (e.g. numbers of clients seen), and subsequently into outcomes that matter to clients impacted by EMD coaching (e.g. improved mental wellbeing). Fifty participants will be recruited including clients who have completed the EMD programme and new clients participating in the online blended learning format. Face-to-face and blended learning formats will offer six sessions over a three-month period. A mixed-method approach will collect quantitative and qualitative from questionnaires and interviews. Outcomes will measure improvements in mental wellbeing and self-efficacy. Wellbeing valuation will be applied to quantify and value outcomes. Social value ratios will be generated from two value sets, one from the Social Value Bank and the other from the Short Warwick Edinburgh Mental Wellbeing Scale.

**Ethics and dissemination:** Ethical approval of this proposed study has been granted by the Medical and Health Sciences ethics committee, Bangor University. A final report will be presented to key stakeholders and research findings will be published in peer reviewed journals

**Article Summary:** *Strengths and limitations of this study:*

- The first study to undertake a SROI analysis on lifestyle coaching to improve mental wellbeing resilience
- Applies a mixed-method approach using both quantitative and qualitative data, using valid and reliable outcome measures
- Researchers working with the same data may arrive at different SROI ratios
- The matching of outcomes from proposed study data with the most appropriate SVB value will depend on the research team’s discretion and agreement. This could potentially introduce researcher bias and the likelihood that potential estimates of social value generated could be upward-biased

## Introduction

In the UK, one in four adults experience poor mental health in their lifetime [1]. Approximately around 1-in-8 adults in the UK are receiving treatments for a diagnosed mental illnesses [2]. During the COVID-19 pandemic, research indicated that 1 in 6 people who required support for a mental health condition could not access mental health services [3]. This evidence highlights current and pressing concerns regarding mental health problems attributed to the social consequences linked with the COVID-19 pandemic [4].

Poor mental wellbeing has the potential for adverse effects on quality of life, relationships, families, corporations, communities and wider society [5]. In addition, poor mental wellbeing impacts on a person’s ability to cope with life, make informed decisions to adequately manage their health and social status [6]. Mental ill health can have a significant impact on life expectancy and is a key cause of health inequalities [7]. Evidence indicates that individuals with severe and enduring mental health problems die on average ten years earlier than the general population [8]. Increasing rates of poor mental health and wellbeing is clearly evidenced in the ongoing crisis of accessing statutory services and the growing prominence of voluntary sector organisations supporting adults and children with mental health challenges [9]. Access to non-medical interventions via the voluntary sector, however, remains challenging for individuals living in isolated or deprived environments with limited support networks and for individuals with who have difficulties accessing statutory services. During the COVID 19 lockdown restrictions in the UK, there was a clear need for innovative, accessible digital interventions to support wellbeing for individuals affected by the pandemic without having to wait many weeks or months on statutory service waiting lists [10].

Social Prescribing (SP) is recognised as an non-clinical intervention that can support mental health services and primary care networks in dealing with individuals living with mental health conditions [11]. EMD is a social prescribing referral option by a Primary Care Cluster in South Wales, UK. SP offers an individual approach to addressing health needs taking account of social, economic and environmental factors, to address needs in a holistic way, and to support individuals to take greater control of their own health [12]. EMD is optimally suited for individuals who are experiencing mental wellbeing challenges, particularly individuals who fall outside statutory mental health service referral criteria and for discharged patients seeking on-going self-development and personal growth.

## Intervention

Prior to the COVID-19 global pandemic, the EMD lifestyle coaching programme was delivered face-to-face. However, because of lockdown restrictions, EMD was adapted and delivered via an online blended format, which featured both live 1:1 support and online content. EMD participants were able to engage with six online modules consisting of 31 learning units covering emotional resilience over a three-month period. The evidence base of the influence of lifestyle coaching has on mental health is limited and requires further exploration [13]. Previous studies have identified that lifestyle coaching has positive effects on reducing psychological stress [15], reducing the likelihood of burnout (Dyrbye *et al*., 2019), along with reducing levels of depression and increasing resilience and confidence (Green, Oades and Grant, 2006). In order to improve the knowledge base, this study applies SROI methodology to evaluate EMD coaching and to estimate the social value gained by participants.

This study is the first evaluation to estimate the social value and effectiveness of lifestyle coaching on improving mental health and wellbeing for people experiencing anxiety and depression.

### Aim

This study will explore the social value generated from EMD coaching as measured by the increase in personal wellbeing experienced by participants.

## Methods

### Study Design

This study will employ a Social Return on Investment (SROI) methodology that includes a mixed-methods approach (questionnaire and semi-structured interview design). Following referral to the EMD programme, participants will receive online forms; consent form, participant information sheet and baseline questionnaire. The baseline questionnaire will capture participant demographic information, reason for referral, current health state, along with information regarding current levels of mental wellbeing and self-efficacy.

The aim of the SROI analysis is to develop a programme-level theory of change to establish how inputs (e.g. costs, staffing) are converted into outputs (e.g. numbers of participants seen), and subsequently into outcomes that matter to participants experiencing the EMD coaching programme (e.g. improved mental wellbeing). The social value generated will then be estimated in a similar way to cost-benefit analysis, and the ratio of social value generated per £1 invested will be calculated. The SROI analysis will be operationalised through the six stages outlined in the Cabinet Office Guide to Social Return on Investment analysis [17]:

1. Identifying stakeholders
2. Developing a theory of change
3. Calculating inputs
4. Evidencing and valuing outcomes
5. Establishing impact
6. Estimating the SROI ratio

### Identifying stakeholders

Stakeholders are defined as individuals or organisations that experience change as a result of the activity being analysed (Cottrel*l et al*, 2014). Stakeholder involvement is an important component of SROI evaluation.

In this study, EMD staff and Bangor University researchers determined that participants who undertook the EMD programme were the primary stakeholder group. Due to the scope of this study, data from other possible stakeholders who may have also benefited from EMD such as family members of participants will not be collected.

### Inclusion criteria

Participant eligibility will include adults (aged over 18 years old) who meet the following criteria:

- Referred to a relevant EMD programme, which lasted approximately three months.
- Participants that are experiencing a physical, mental or social issue that would benefit from the EMD intervention.
- Participants that have the mental capacity to reflect on their own wellbeing.
- Participants who speak Welsh or English

Recruitment will be conducted by means of local university health boards, social media sites, through social care and health care organisations as well as by way of the national Social Prescribing Network.

### Patient and Public Involvement

The research question (i.e., ‘what is the social return on investment of the EMD programme’) was informed by client experience and feedback on the EMD programme. In 2019, 24 previous clients took part in the EMD programme and completed an informal online questionnaire, which indicated substantial improvements in mental wellbeing, self-confidence and empowerment. These client responses helped inform the outcome measures used in the current SROI study.

Although client feedback helped to inform the choice of outcome measures in the SROI study, clients were not involved in the actual design of the SROI study.

Previous face-to-face clients and new online clients were recruited into the SROI study. All clients completed questionnaires and interviews which provided both quantitative and qualitative data used in this SROI study.

Study participants will be informed by email when the final report is completed. Study participants will be provided with a link to view the final report which will be available on the EMD web page, the Social Value Hub, Bangor University web page as well Cardiff University Clinical Innovation Hub and News webpages. In addition, SROI results will be disseminated to a wider audience through the community of practice through the Wales School for Social Prescribing Research (WSSPR).

### Data Collection and Management

Questionnaire data and consent forms will be stored on an online questionnaire platform. Only authorised personnel will have access to the data. Additional security measures for the platform include continual monitoring of up-time (when the platform is live and accessed online) as well as unauthorised attempts for admin logins. Regular back-ups of the entire website are automated, retaining a copy of the website files for emergency use. The website is hosted on UK based hosted servers which have 24/7 support where required. Responses from the questionnaires will be manually entered into Microsoft Excel for summary and analysis.

Interview audio recordings and transcriptions will be uploaded to a secure network that only core members of the advisory team will have access to.

### Developing a theory of change

The purpose of the theory of change model is to develop and determine the causality and impact of an intervention on the participants [19]. Informed by a rapid review of literature along with existing informal evidence from EMD clients, a theory of change will be generated to explain the relationship between the programme inputs, outputs and outcomes for EMD stakeholders, and how these outcomes could generate social value.

### Calculating inputs

The SROI protocol will examine the total costs for the EMD coaching programme that will include both administration costs and coaching session costs. Administration costs involve the time for EMD staff to monitor and evaluate existing programmes as well as to coordinate and develop referrals. Session costs involve minimum and maximum cost scenarios for one EMD coach and coaching materials. After the quantity of outcomes has been determined for face-to-face and online clients via the questionnaires and interviews, monetary values will be assigned to mental wellbeing and self-efficacy outcomes using sources such as the HACT Social Value Bank (https://www.hact.org.uk/social-value-bank), Social Value Calculator (https://www.hact.org.uk/value-calculator) and Mental Health Social Value Calculator (https://www.hact.org.uk/mental-health-social-value-calculator. To estimate the value of the quantity of change in health service use for mental health conditions, the Curtis and Burns Unit Costs of Health and Social Care (2021) and NHS Reference Costs (2021) will be applied to calculate the programme inputs.

### Evidencing and valuing outcomes

#### Questionnaires

Data from the new online participants will be collected via pre and post intervention questionnaires. Previous face-to-face clients will complete a one-time only questionnaire to estimate the changes clients experienced in mental wellbeing and self-efficacy during the time they participated in the EMD programme.

Questionnaires will collect information about participant demographics (gender, age, ethnicity, occupation, and location), and reasons for undertaking the EMD coaching programme. In addition, information will be collected on participants, mental wellbeing, and self-efficacy. Data on engagement with mental health services will compare health service usage three months prior to commencing the EMD coaching programme along with the three months during the programme.

Mental wellbeing will be measured by means of the Short Warwick-Edinburgh Mental Wellbeing Scale (SWEMWBS), which is a list of seven positively worded statements with five coded response categories to measure different aspects of positive mental health [20]. Overall scores can range from 7 to 35 and the SWEMWBS has a good construct validity and good test retest reliability [21].

Self-efficacy will be assessed with the General Self-Efficacy Scale (GSES) which is a 10-item self-report measure of self-efficacy range. This scale assesses the strength of an individual’s belief in their ability to respond to novel or difficult situations as well as to deal with any associated obstacles or setbacks (Schwarzer and Jerusalem, 1995). Overall scores can range from 10 to 40 and the GSES range is a reliable and valid when measuring self-efficacy and confidence, as the ten items outlined are relevant to self-efficacy amongst individuals [22].

A Client Service Receipt Inventory (CSRI) questionnaire, which will be employed to estimate health service use for mental health-related conditions. The CSRI will allow participants to report how many times the following services were accessed three months prior to commencing the EMD coaching programme and three months during the EMD programme. These services include:

- Number of appointments with a Psychiatrist
- Number of appointments with a Psychologist
- Number of appointments with a Mental Health Nurse
- Number of appointments with a Social Worker
- Number of appointments with any other community based mental health service

#### Interviews

Participants will be invited to attend a semi-structured interview. The qualitative interviews will explore participants’ lived experience of participating in the EMD coaching programme. Informed consent will be obtained from participants prior to the being conducted by phone or Zoom. All interviews will be audio-recorded and transcribed.

#### Social Value Analysis

Social value analysis is a means of assessing the impact of interventions on wellbeing by estimating the social value generated for stakeholders [23]. In this study, wellbeing valuation will be applied and offers a consistent and robust method for estimating the monetary value of outcomes that do not have market values. Two value sets will be used: the Social Value Bank (SVB) and SWEMWBS value set.

SVB will be used to monetise the outcomes of increased self-efficacy and the SWEMWBS value set will be used to monetise mental wellbeing. Since, the values in the SVB incorporate mental wellbeing, the two value sets (SVB and SWEMWBS) are treated separately rather than as two value sets that can be combined [24,25].

### Establishing impact

To establish impact, it is necessary to reduce bias and decrease the risk of over-claiming the benefits of EMD coaching programme. The follow-up questionnaires and interviews will include specific questions to determine deadweight (the proportion of observed outcomes that may have happened anyway); displacement (the proportion of outcomes that may have been displaced from one sector to another); attribution (the amount of outcome directly attributed to EMD coaching); and drop-off (the length of time that outcomes last for EMD clients) [26].

### Estimating the SROI ratio

The SROI ratio will be calculated by dividing the total value of inputs by the total value of outcomes. The proposed resulting ratio is the amount of social value generated for every £1 invested in EMD coaching programme as outlined in Figure 1. In addition to calculating a proposed base case scenario, a range of sensitivity analyses will be performed to explore how the SROI ratio could be affected if different financial proxies were used and if varying levels of outcomes were achieved than those used in the base case scenario. A checklist for quality assessment in SROI analysis will be used as a framework to guide the reporting of the SROI findings (Hutchinson *et al*, 2018).

**Figure 1:**
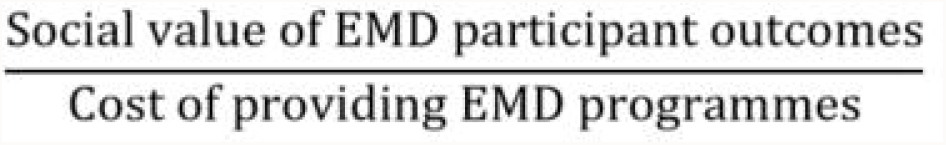
Social Return on Investment Ratio equation.

## Discussion

This study will determine the social value generated from the EMD lifestyle coaching programme and compare this with the costs of delivering the programme to generate a SROI ratio. The effectiveness of the EMD programme will be ascertained by comparing baseline and follow-up mental wellbeing and self-efficacy scores and CSRI data.

The effectiveness of the EMD programme will be compared with existing literature evaluating other lifestyle coaching programmes. For example, Aboalshamat et al (2020), found that lifestyle coaching had a positive impact on stress among dental students (n=88). Green, Oades and Grant (2006) reported positive effects of lifestyle coaching on a non-clinical population (n=56). Dyrbye et al (2019) found that lifestyle coaching reduced the likelihood of burnout among physicians (n=88)

### Advisory Team

The advisory team for this study will comprise of the EMD Coach, researchers from Bangor University, Cardiff University, and the Hywel Dda University Health Board. The advisory team will meet weekly for meetings to discuss recruitment, data collection progress, and the write-up of the stakeholder report.

### Ethics and dissemination

Ethical approval of this study has been approved by Medical and Health Sciences ethics committee, Bangor University (Reference number: 2021-16927). Participant data will be anonymised, and no participants will be identifiable in the results. Participants will be reminded that they can withdraw from the study at any time without explanation.

A full report will be submitted to the funders at the conclusion of the study and findings will be available to the key stakeholders and the general public. In addition, academic papers will be submitted for publication to peer-reviewed academic journals.

## Conclusion

This novel SROI study will determine whether the EMD lifestyle coaching programmes, face-to-face format and online blended learning format, have the potential to generate positive social value ratios. Furthermore, this study will compare the effectiveness of the two lifestyle coaching formats: face to face and the online blended format to improve mental wellbeing. Finally, the proposed mixed-method approach applied within the SROI methodology will indicate whether participants experience improvements in mental wellbeing and self-efficacy by participating in this innovative lifestyle coaching programme.

## Data Availability

Questionnaire data and consent forms will be stored on an online questionnaire platform. Only authorised personnel will have access to the data. Additional security measures for the platform include continual monitoring of up-time (when the platform is live and accessed online) as well as unauthorised attempts for admin logins. Regular back-ups of the entire website are automated, retaining a copy of the website files for emergency use. The website is hosted on UK based hosted servers which have 24/7 support where required. Responses from the questionnaires will be manually entered into Microsoft Excel for summary and analysis.
Interview audio recordings and transcriptions will be uploaded to a secure network that only core members of the advisory team will have access to.
The final dataset will only be available to the study investigators and the Advisory Team.

## Contributors

The study concept and design was conceived by ML, AM, NH, AC, EW, and RTE. NH, EW and AM will conduct screening and data collection. Analysis will be performed by AM, NH, and ML. ML, NH, AM, and AC prepared the first draft of the manuscript. All authors provided edits and critiqued the manuscript.

## Competing Interest

None declared.

## Funder

Accelerate: the Welsh Health Innovation and Technology Accelerator East Wales ERDF No. 80751.

## Data Sharing

The final dataset will only be available to the study investigators and the Advisory Team.

## Acknowledgements

We would like to acknowledge Hayley T Wheeler, the founder and lead Practioner of the EmotionMind Dynamic. This research would not be possible without your support, knowledge, enthusiasm and commitment to Public Mental Health and Wellbeing.

## Notes

### Competing Interest Statement

The authors have declared no competing interest.

### Funding Statement

This study was funded by Accelerate: the Welsh Health Innovation and Technology Accelerator
East Wales ERDF No. 80751.

### Author Declarations

Ethics and dissemination Ethical approval of this study has been approved by Medical and Health Sciences ethics committee, Bangor University (Reference number: 2021-16927). Participant data will be anonymised, and no participants will be identifiable in the results. Participants will be reminded that they can withdraw from the study at any time without explanation. A full report will be submitted to the funders at the conclusion of the study and findings will be available to the key stakeholders and the general public. In addition, academic papers will be submitted for publication to peer-reviewed academic journals.

